# Half of children entitled to free school meals do not have access to the scheme during the COVID-19 lockdown in the UK

**DOI:** 10.1101/2020.06.19.20135392

**Authors:** Jennie C Parnham, Anthony A Laverty, Azeem Majeed, Eszter P Vamos

## Abstract

**Objectives:** To investigate access to free school meals (FSM) among eligible children, to describe factors associated with uptake and investigate whether receiving FSM was associated with measures of food insecurity in the UK using the COVID-19 wave of the UK Household Longitudinal Study (UKHKS).

**Study design:** Cross sectional analyses of questionnaire data collected in April 2020.

**Methods:** 635 children who were FSM eligible with complete data were included in the analytic sample. Accessing a FSM was defined as receiving a FSM voucher or a cooked meal at school. Multivariable logistic regression was used to investigate (i) associations between characteristics and access to FSM and (ii) associations between access to FSM and household food insecurity measures. All analyses accounted for survey design and sample weights to ensure representativeness

**Results:** 51% of eligible children accessed a FSM. Children in junior schools or above (aged 8+ years) (Adjusted Odds Ratio (AOR) 11.81; 95% CI 5.54,25.19), who were low income (AOR 4.81; 95% CI 2.10,11.03) or still attending schools (AOR 5.87; 95% CI 1.70,20.25) were more likely to receive FSM. Children in Wales were less likely to access FSM than those in England (AOR 0.11; 95% CI 0.03,0.43). Receiving a FSM was associated with an increased odds of recently using a food bank, but not reporting feeling hungry.

**Conclusions:** In the month following the COVID-19 lockdown, 49% of eligible children did not receive any form of FSM. The present analyses highlight that the voucher scheme did not adequately serve children who could not attend school during the lockdown. Moreover, more needs to be done to support families relying on income-related benefits, who still report needing to access a foodbank. As the scheme may be continued in summer or in a potential second wave, large improvements will be needed to improve its reach.

## INTRODUCTION

In the UK, 10% of children experience severe food insecurity with research suggesting levels have risen during the COVID-19 pandemic^1,2^. As food insecurity is associated with a wide range of negative health outcomes, including increased hospitalisations, asthma and poor mental health^3^, it is critical that food insecurity prevalence does not increase. Free school meals (FSM) are a key public health policy in reducing food insecurity and dietary inequalities in children in the UK. Currently there are two FSM schemes in the UK. Under the means-tested scheme, children in a household receiving income-related benefits are eligible for a FSM. As of 2014 however, a universal FSM scheme was introduced, in which all infant school children (aged 4–7 years) are eligible for a free meal, regardless of income. Research has shown that school meals play an important role in levelling inequalities in dietary intake, with the most deprived children having the most to gain from eating a school lunch^4,5^. Moreover, school lunches have been shown to be healthier than packed lunches regardless of income-level^6^. As such, the universal infant FSM scheme has been associated with reduced obesity rates in Reception (ages 4–5 years) ^7^.

On 20^th^ March 2020, all UK schools closed until further notice due to COVID-19, except to vulnerable children and children of key workers. Consequently, the 1.3 million children who claim FSM in England were unable to access their entitlement unless they were eligible to attend a ‘skeleton’ school (were vulnerable or a child of a key worker)^8^. Vouchers worth £15 per week were introduced from 31^st^ March, to ensure FSM eligible children had continued access to lunch outside of school. Schools had the responsibility of applying for and distributing electronic voucher codes to their free school meal eligible pupils. The scheme was only made available to children on the means-tested FSM scheme, not the universal infant FSM scheme. Since implementation, there have been reports some beneficiaries were still not able to access FSM^2^. In a time of sudden economic change, which has been shown to affect those on low-incomes the worst^9^, it essential the government ensures continuity of the FSM scheme in the COVID-19 lockdown. Optimal implementation of the scheme will be needed to support low-income children and prevent a widening of health inequalities.

## OBJECTIVES

We investigated access to free school meals (FSM) among eligible school children in the UK using the COVID-19-wave of the UKHLS. Additionally, we described factors associated with uptake and investigated whether receiving FSM was associated with measures of food insecurity.

## METHODS

UK Household Longitudinal Survey (UKHLS) participants were invited to answer a COVID-19 questionnaire between 17^th^ and 30^th^ April 2020. A child-level dataset was produced from the proxy-responses of a guardian in the household (n=4,559). The analytic sample included 635 children who had complete data and self-reported as FSM eligible. FSM eligibility did not distinguish between means-tested and universal schemes. Accessing FSM was defined as having received a FSM voucher or a cooked meal at school. Logistic regression was used to investigate (i) associations between characteristics and access to FSM and (ii) associations between access to FSM and household food insecurity measures. Characteristics in the model included: school phase; ethnicity of guardian, household income, country and school attendance during lockdown. Household income was taken from wave 9 of UKHLS (2017-19) as the variable was more complete and could be equivalised for household composition (OECD scale)^10^. Participants with missing income information were included in a fourth category. Measures of household food insecurity include reporting using a food bank in the last four weeks and reporting a household member feeling hungry but being unable to eat in the past week. All analyses accounted for survey design and sample weights to account for non-response and make the results representative to the UK population.

## RESULTS

In the analytic sample, 635 children reported being eligible for FSM, 49% of whom did not receive any form of FSM entitlement in April 2020 (see Table 1). Our analyses found that children who were in the lowest income category were almost five times more likely to receive their FSM entitlement than high income children (OR 4.81; 95% CI 2.10,11.03). Children who were still attending school, were almost six times more likely to receive their FSM entitlement than children who could not (OR 5.87; 95% CI 1.70,20.25). Children in Wales, compared to England, were 89% less likely to access a FSM (OR 0.11; 95% CI 0.03,0.43). The analyses showed a large difference in the odds of receiving a FSM between school phase. Those in junior and secondary schools were more likely to access FSM than those in infant schools (OR 11.81 and 16.45, respectively). An interaction between income-level and school phase was tested to investigate whether the association between income and receiving FSM differs by school phase of child. The interaction term was not statistically significant.

**Table 1.** Characteristics associated with receiving a free school meal among children in lockdown among who are eligible for a free school meal in April 2020.

|  | Did the child access their free school meal? |  |  |  |  | Logistic regression <sup>‡</sup> |  |
| --- | --- | --- | --- | --- | --- | --- | --- |
|  | No<br>(n=341, 49%) |  | Yes<br>(n=294, 51%) |  | Total<br>(n=635) |  |  |
|  | *n* | (%) | *n* | (%) | *n* | OR | 95% CI |
| **School phase** |  |  |  |  |  |  |  |
| *Infants (ages 4-7)* | 284 | (77.26) | 75 | (22.74) | 359 | *ref* |  |
| *Juniors (ages 8-11)* | 30 | (23.69) | 93 | (76.31) | 123 | 11.81*** | [5.54,25.19] |
| *Secondary (ages 12-18)* | 27 | (16.94) | 126 | (83.06) | 153 | 16.45*** | [7.59,35.66] |
| **Guardian's Ethnicity** |  |  |  |  |  |  |  |
| *White* | 315 | (49.75) | 252 | (50.25) | 567 | *ref* |  |
| *BAME* | 63 | (43.49) | 80 | (56.51) | 143 | 0.65 | [0.09,4.82] |
| **Equivalised household income <sup>†</sup>** |  |  |  |  |  |  |  |
| *Low* | 123 | (35.02) | 192 | (64.98) | 315 | 4.81*** | [2.10,11.03] |
| *Middle* | 119 | (61.84) | 79 | (38.16) | 198 | 2.46 | [1.00,6.10] |
| *High* | 99 | (81.94) | 23 | (18.06) | 122 | *ref* |  |
| *Missing* | 37 | (46.54) | 38 | (53.46) | 75 | 1.9 | [0.72,5.02] |
| **Country** |  |  |  |  |  |  |  |
| *England* | 321 | (46.73) | 283 | (53.27) | 604 | *ref* |  |
| *Wales* | 13 | (75.75) | 17 | (24.25) | 30 | 0.11** | [0.03,0.43] |
| *Scotland* | 36 | (65.59) | 19 | (34.41) | 55 | 0.66 | [0.21,2.05] |
| *Northern Ireland* | 8 | (53.02) | 13 | (46.98) | 21 | 0.23 | [0.01,4.81] |
| **Child at school in lockdown** |  |  |  |  |  |  |  |
| *Yes* | 16 | (21.49) | 32 | (78.51) | 48 | 5.87** | [1.70,20.25] |
| *No* | 362 | (51.23) | 300 | (48.77) | 662 | *ref* |  |

\* $P<0.05$ ; \*\* $P<0.01$ ; \*\*\* $P<0.05$ . *ref* = reference group. OR = odds ratio. CI = confidence interval.
BAME = Black and minority ethnic
<sup>†</sup> Wave 9 household income (2017-18) equivalised using OECD scale and categorised into quantiles
<sup>‡</sup> Multivariable logistic regression with school phase, Guardian's ethnicity, household income, country and school attendance included in the model.

In a second multivariable logistic regression model which controlled for the same characteristics, we assessed whether access to FSM was associated with measures of food insecurity. First, access to FSM was not associated with someone in the household feeling hungry but being unable to eat in the past week (OR 0.99; 95% CI 0.35,2.82). Second, those who accessed their FSM entitlement were found to be 14 times more likely to have recently used a foodbank (OR 13.89; 95% CI 2.27,85.10).

## DISCUSSION

The present analyses demonstrate that a significant proportion of eligible children could not access free school meals during the COVID-19 lockdown. As children who attended school were more likely to receive a meal, the results indicate that the FSM vouchers did not act as a sufficient replacement for receiving a meal at school.

These data also imply that pupils at secondary schools had better access to some form of the FSM scheme than pupils at infant and junior schools. However, the assessment of FSM eligibility in the study did not distinguish between the means-tested and universal scheme. Consequently, infant school children on the universal scheme but not eligible for the means-tested scheme may be misclassified. If the results were predominantly due to misclassification, we would expect to see an effect modification by income-level, which was not apparent as the interaction term was insignificant. Among FSM eligible children, the lowest-income children were more likely to access FSM. Low-income households have been most greatly impacted by the COVID-19 lockdown^9^, so higher uptake likely reflects a greater need in these households to limit food insecurity. This hypothesis is supported by the increased likelihood of foodbank use among children who accessed FSM. Use of foodbanks in this group reveals an inadequacy of government welfare schemes to protect vulnerable, low-income families in the UK from food insecurity.

## CONCLUSION

In the first month which UK schools were closed by COVID-19, this study used nationally representative data to highlight that half of all eligible children did not receive FSM. It is currently unclear when all pupils will return to school. As such, it is concerning that children from low-income families who cannot attend schools may continue to not have access to nutritious meals, putting their physical and mental health at risk. This issue will only grow more critical in upcoming school summer holiday. Without increased intervention and support, low-income families are at risk of increased food insecurity and negative health consequences during the COVID-19 pandemic, which will likely be exacerbated by the economic recession which will follow.

## Data Availability

Data is available to download from UK data service (Study Number 8644)

https://beta.ukdataservice.ac.uk/datacatalogue/studies/study?id=8644

